# The genetic determinants of plasma protein variance across ancestries and effects on cardiometabolic disease risk

**DOI:** 10.1101/2025.11.20.25340688

**Authors:** Chief Ben-Eghan, Elodie Persyn, Carles Foguet, Zongtai Wu, Xilin Jiang, Yu Xu, Scott Ritchie, Samuel A. Lambert, Adam S. Butterworth, Stephen Burgess, Michael Inouye

**Affiliations:** Cambridge Baker Systems Genomics Initiative, Department of Public Health and Primary Care, University of Cambridge, Cambridge, UK; British Heart Foundation Cardiovascular Epidemiology Unit, Department of Public Health and Primary Care, University of Cambridge, Cambridge, UK; Victor Phillip Dahdaleh Heart and Lung Research Institute, University of Cambridge, Cambridge, UK; British Heart Foundation Centre of Research Excellence, University of Cambridge, Cambridge, UK; Health Data Research UK Cambridge, Wellcome Genome Campus and University of Cambridge, Cambridge, UK; Cambridge Baker Systems Genomics Initiative, Baker Heart and Diabetes Institute, Melbourne, Victoria, Australia; MRC Biostatistics Unit, University of Cambridge, Cambridge, UK

## Abstract

Variance quantitative trait loci (vQTLs), which capture genetic contributions to phenotypic variability, remain underexplored in proteomic studies, particularly across diverse ancestries. We systematically mapped *cis*-vQTLs for 2,923 plasma proteins in 52,706 UK Biobank participants of European (EUR, *N* = 45,486), African (AFR, *N* = 1,336), and Central/South Asian (CSA, *N* = 934) ancestries, identifying 2,162 vQTLs (P_VE_ < 5 x 10^-8^) for 781 proteins. We identified ancestry-specific and shared *cis*-vQTLs, including those for 30 proteins which were shared across all ancestries, with a few proteins, exhibiting stronger associations in non-EUR ancestry groups despite smaller sample sizes. Across ancestries, 7% (EUR), 25% (AFR), and 14% (CSA) of associations had variance effects only (vQTL_only_), lacking corresponding mean effects (P_ME_ > 0.05), with chromosome X enriched for vQTL_only_ associations. Finally, multivariable Mendelian randomization revealed that, independent of genetically predicted mean protein levels, genetically predicted variance of three proteins influenced disease risk of coronary artery disease (Lp(a) and VAMP5) or type 2 diabetes (ANGPTL4). The MR effects for protein levels and variance were independent yet directionally consistent and significant (FDR < 0.05). Taken together, this study identifies novel protein vQTLs, highlights their transferability and demonstrates the potential therapeutic relevance of protein variance.

## Introduction

The human plasma proteome provides a snapshot of human health^1,2^, and high-throughput proteomic platforms, such as Olink and SomaLogic, have enabled interrogation of the proteome across large cohorts of individuals at scale^3–5^. Despite rapid progress in omics technologies, high costs remain a major barrier to widespread adoption. Expanding proteomic studies to more diverse populations is, however, essential to better reflect global genetic and proteomic variation, and ensure findings are more equitable and relevant to a broader range of individuals^6^.

Proteomic profiles are highly dynamic, varying spatiotemporally across individuals, populations and ancestries^7,8^ due to complex genetic and environmental interactions which are poorly understood^9^. Most genome-wide proteomic studies to date have focussed on capturing only additive genetic effects (main effects, based on trait means) associated with the proteome. More recently, variance quantitative trait loci (vQTLs), loci associated with different trait variances across genotypes, have been characterized in complex traits such as body mass index (BMI) and blood cell traits^9,10^. vQTLs can also serve as proxies for gene-environment interactions (GEI) and provide insights into the role genetics plays in influencing phenotypic plasticity^9^. Specifically, studies typically evaluate vQTLs as modulating the variance of protein levels across individuals within populations, rather than capturing variability from longitudinal or repeated-measures for a single individual. For example, a recent study that characterized vQTLs using the initial data release of ∼1,500 plasma proteins from the UK Biobank (UKB), highlighted the enrichment of vQTLs in GEI associations for participants of European ancestries^11^.

There is mounting evidence that the likelihood of success in developing a drug against an actionable target is at least 2-fold higher when supported by human genetic evidence^12^. This is particularly evident when considering proteins as targets, since an effective drug frequently alters protein function by modulating its levels, structure or activity, thereby affecting disease risk. To date, Mendelian randomization (MR) has been useful in establishing causal links between proteins and several complex traits by leveraging genetic variants as instrumental variables to simulate naturally occurring randomized trials^13,14^. For example, MR studies have shown a linear dose-response relationship between lipoprotein(a) (Lp(a)) and coronary artery disease (CAD) risk, with greater risk reduction at higher Lp(a) levels. In one study, the highest efficacy for inhibition of Lp(a), was detected at levels >70 mg/dL, which directly influenced the design of Novartis’ Phase III cardiovascular trial ‘HORIZON’ ^15,16^. *Cis*-MR leverages the additive effects of genetic variants proximal to a protein-coding gene, to infer the causal role of the encoded protein on a specific outcome. However, MR studies rarely model both genetically predicted protein levels and genetically predicted protein variance as exposures, thus the causal role of protein variance for common diseases is largely unknown. Multivariable MR (MVMR) represents a potential solution by extending the classic MR framework to simultaneously model multiple exposures against an outcome^17^.

This study aims to address critical gaps in our knowledge of protein vQTLs. Here, we use UK Biobank to extend our knowledge of *cis*-vQTL mapping to 2,923 proteins in European, African and Central/South Asian ancestries, then assess the putatively causal role of protein variance in disease. In doing so, we characterize ancestry-specific and shared vQTLs and conducted GEI analyses for five common demographic and behavioural factors as exposures. Multivariable MVMR is then used to quantify the distinct effects of protein levels and protein variances on risk of coronary artery disease (CAD) and type 2 diabetes (T2D), thus informing drug target prioritisation. Taken together, our results suggest the potential for protein variances to inform therapeutic development strategies.

## Results

### Characterising vQTLs across diverse ancestries

To define ancestry groups within the UKB participants with Olink Explore 3072 proteomic data^3^ (*N* = 52,706; 2,923 proteins), we used 1000 Genomes (1KG) continental superpopulations as reference for genetic similarity analysis, identifying three groups with sufficient sample sizes for vQTL analyses: African (1KG-AFR-like, *N* = 1,336), European (1KG-EUR-like, *N* = 45,486), and Central/South Asian (1KG-CSA-like, *N* = 934) (**Methods**; **Figure S1**; **Table S1**). For brevity, population descriptors are abbreviated as three-letter codes (AFR, EUR, and CSA respectively) and are used to indicate how the data were analysed statistically (**Methods**). Importantly, while statistically useful, these clusters do not reflect the full continuum of genetic variation in these geographical locations and do not reflect cultural or ethnic groups^18^. Given the substantially larger sample size, we present the EUR results first and then follow with non-EUR ancestries for comparison. For clarity, we denote variance effects with subscript _VE_ and mean effects with _ME_.

Across all ancestries, we identified a total of 2,162 near-independent *cis-*vQTLs (vQTLs within ±1Mb of the transcription start sites (TSS)) at a significance threshold of P_VE_ < 5×10^-^⁸ (**Methods**) with, each vQTL associated with at least one of 781 proteins. These included 2,001 vQTLs (771 proteins) detected in EUR; 104 vQTLs (84 proteins) in AFR and 61 vQTLs (54 proteins) in CSA (**Table S2**). As expected, the majority of vQTLs (92.3%) were detected in EUR, driven by its greater statistical power for discovery (**Figure S2**), with associations in AFR and CSA representing 4.8% and 2.9% of significant vQTLs, respectively (**Figure 1a**). Of proteins with a detected vQTL, approximately half had more than one significant, near-independent vQTL in EUR (EUR 432 proteins, 56.0%; AFR 13 proteins, 15.5%; CSA 5 proteins, 9.3%). In EUR, as many as 15 near-independent vQTLs were detected for a single protein, IL13RA1 (**Table S2**). In AFR, XPNPEP2 had the highest count with seven near-independent vQTLs, and in CSA, CD200R1 showed the highest count with three (**Figure 1a; Table S2**).

**Figure 1:**
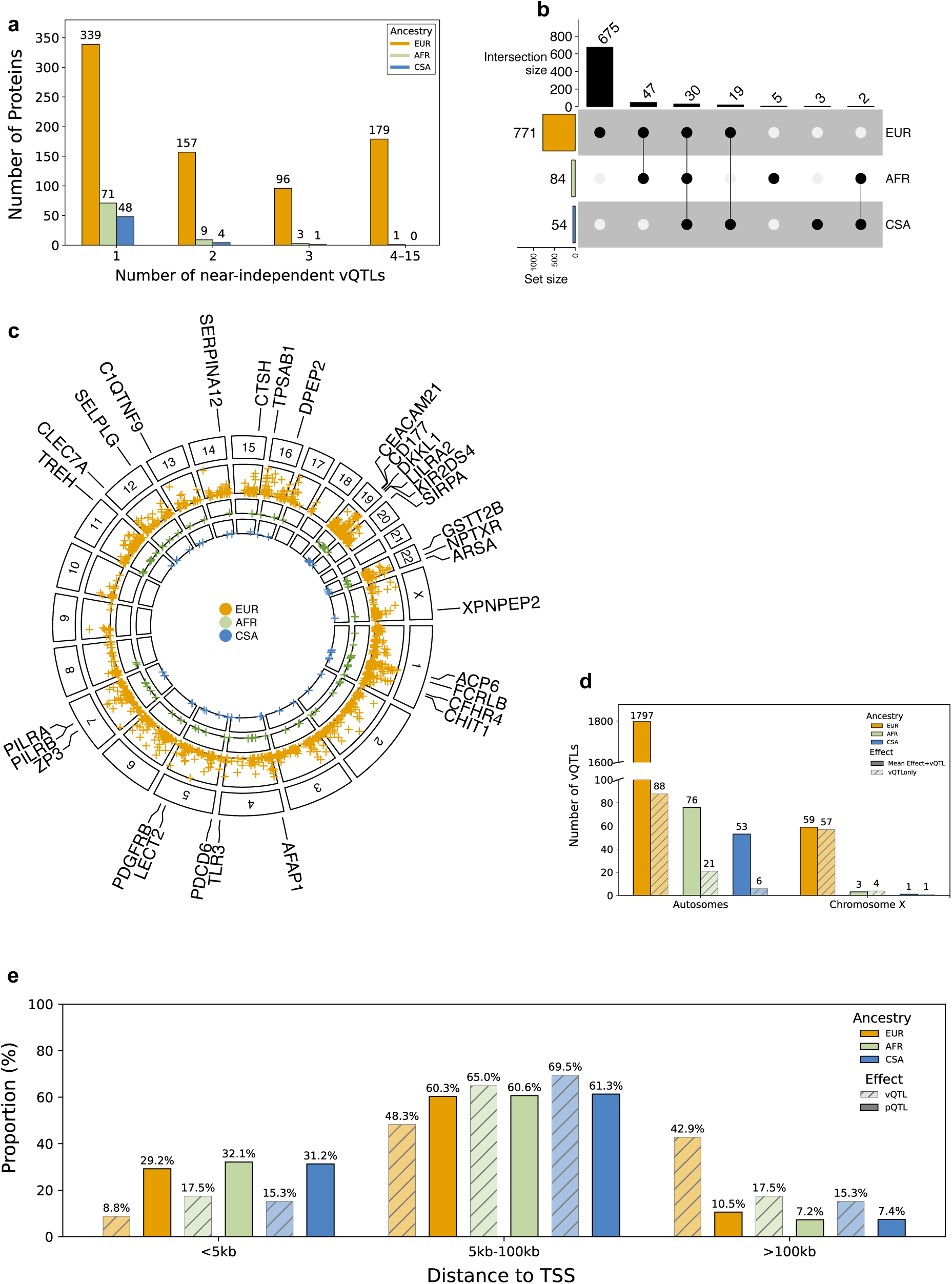
Summary of vQTL associations identified across multiple ancestries. Panel (**a**) shows the number of near-independent vQTLs identified per protein across ancestries with the total number of proteins labelled above each bar. Panel (**b**) presents an UpSet plot illustrating the union of all vQTLs, categorized as ancestry-specific or ancestry-shared based on the overlap of proteins. The bottom panel uses black dots and connecting lines to indicate proteins with ancestry-shared vQTL associations, while black isolated dots represent proteins with only ancestry-specific vQTLs. The bars above and to the left of the plot represent the number of proteins contributing to each combination of ancestries and individual ancestry totals, respectively. Panel (**c)** is a Circos Manhattan plot highlighting 30 proteins with vQTLs shared across all three ancestries. Panel (**d**) separates identified vQTLs into those located on autosomes and those on chromosome X. Panel (**e**) shows the proportion of cis-vQTLs and *cis*-pQTLs from Sun et al^3^, across three distances to TSS bins, stratified by ancestry.

We identified 30 proteins (29 autosomal, one coded on chromosome X) which shared vQTLs across all three ancestries (EUR: 165 vQTLs; AFR: 32 vQTLs; CSA: 31vQTLs) (**Figure 1b-c)**. Most vQTLs (92%) exhibited independent mean effects on the same protein (at least P_ME_ < 0.05), consistent with previous research^11^. Within each ancestry, a subset of vQTLs were identified with vQTL only effects (vQTL_only_), lacking a statistically significant mean effect (P_ME_ > 0.05). The distribution of vQTLs with vQTL_only_ effects by ancestry for autosomes was: 88 vQTL_only_ in EUR (7.3%), 21 vQTL_only_ in AFR (24.5%), and 6 vQTL_only_ (12.9%) in CSA (**Figure 1d, Table S2**).

We also performed a separate analysis on chromosome X, excluding pseudo-autosomal regions, which revealed a significant enrichment of vQTL_only_ effects compared to autosomes (**Methods, Figure 1d, Table S2**). In EUR, 117 vQTLs were detected on chromosome X, of which 58 (49.5%) exhibited vQTL_only_ effects. In AFR, 4 of 7 vQTLs, while in CSA, 1 of 2 vQTLs exhibited vQTL_only_ effects (**Figure 1d)**. However, findings in AFR and CSA should be interpreted with caution given the relatively small number of independent vQTLs compared to the EUR. Notably, only the metalloprotease XPNPEP2 on chromosome X had vQTLs which were shared across all three ancestries (**Table S2**). Finally, we compared the proportion of *cis*-vQTLs and *cis*-pQTLs from Sun et al^3^, across three distances to TSS bins, stratified by ancestry. Across ancestries, ∼50% of both QTLs fell within 5 to 100 kb of their respective TSSs. In the EUR, ∼40% of *cis*-vQTLs were located more than 100 kb from the TSS compared to ∼10% for *cis*-pQTLs. In contrast, only ∼8% of *cis-*vQTLs fell within 5 kb of the TSS, whereas ∼30% of *cis*-pQTLs mapped to this region in the EUR (**Figure 1e**).

We observed similar patterns when we mapped vQTL lead variants to functional annotations **(Figure S3a-c**). Across all ancestries, the majority of annotated vQTLs were consistently located in introns, accounting for 45.3% in EUR (749 vQTLs), 47.9% in AFR (46 vQTLs), and 47.3% in CSA (26 vQTLs); while variants in exons represented a smaller proportion of vQTLs: 3.7% in EUR (64 vQTLs), 11% in AFR (8 vQTLs), and 4.8% in CSA (4 vQTLs) (**Figure S3a-c**). We used ANNOVAR, implemented in FUMA^19^ to assess vQTL enrichment across functional annotation classes against a background of all annotated, ancestry-matched variants in the same regions (**Methods**, **Figure S3d**; **Table S3**). Ranked by both fold enrichment and Fisher’s exact test p-value significance, across all ancestries, the strongest enrichments were observed in 3’ untranslated regions (UTR3; EUR: 2.5-fold, p = 7.8 x 10^-135^; AFR: 3.3-fold, p = 5 x 10^-13^; CSA: 4.1-fold, p = 1.8 x 10^-22^) and exonic regions (EUR: 2.1-fold, p = 1.46 x 10^-81^; AFR: 3.2-fold, p = 2.8 x 10^-13^; CSA: 2.5-fold, p = 2.5 x 10^-8^ (**Figure S3d**). In contrast, intronic regions showed significant depletion (EUR: 1.3-fold, p = 3.1 x 10^-300^; AFR: 1.7-fold, p = 1.6 x 10^-94^; CSA: 1.4-fold, p = 4.7 x 10^-49^) with similar depletion observed in the intergenic regions (EUR: 0.7-fold, p = 1.46 x 10^-300^; AFR: 0.4-fold, p = 2.3 x 10^-118^; CSA: 0.5-fold, p = 4.9 x 10^-80^). Thus, overall, vQTLs were disproportionately enriched in 3’ untranslated regions and coding regions even though greater vQTL density across ancestries was observed in introns and intergenic regions (**Figure S3d**; **Table S3**). Across all ancestries, vQTLs also showed an inverse logarithmic relationship between effect size and minor allele frequency (MAF) with Pearson correlation coefficients of *r =* −0.36 for EUR; −0.68 for AFR, and −0.69 for CSA respectively (**Figure S4a-c**). This pattern is likely influenced by lower statistical power in the non-EUR ancestry groups.

### Cross-ancestry associations

We conducted a systematic, pairwise comparison of independent vQTL effect sizes across all ancestry combinations to assess consistency in terms of directionality and magnitude. To achieve this, for each ancestry pair, we extracted all independent vQTLs identified in the first population and then mapped them to the corresponding variants in the second population (target). Any variant that failed quality control (QC) in the target population was excluded from the comparison (**Methods)**. For robustness of estimates, we restricted our analysis to vQTLs with a MAF > 5% in the target population^9^. When we compared vQTL effect sizes between independent vQTLs from EUR mapped to CSA, we observed a moderate degree of correlation (*r* = 0.69, *p* = 1.5×10^-34^), with approximately 19.5% of EUR vQTLs replicating at nominal significance (P_VE_ < 0.05) and exhibiting consistent effect direction in CSA (**Figure 2a**). In contrast, the correlation (*r* = 0.08, *p* = 0.32) and replication proportions (11.2%) between EUR vQTLs mapped to AFR was much weaker, suggesting limited transferability of vQTL effect sizes from EUR to AFR (**Figure 2b**). Next, we observed strong concordance when mapping AFR effect sizes onto EUR (*r* = 0.8, *p* = 5×10^-3^) and onto CSA (*r* = 0.99, *p* = 6 x10^-7^). Moderate correlation was observed from CSA to EUR (*r* = 0.5, *p* = 0.15) and strong correlation with CSA to AFR (*r* = 0.81, *p* = 0.016) (**Figure S5**). The proportions of replication were higher with non-EUR to EUR pairwise-comparisons. At a nominal threshold of P_VE_ < 0.05, the AFR to EUR replication gave 10 out of 25 vQTLs and the CSA to EUR replication 10 out of 11 vQTLs; at the stricter P_VE_ = 5 x 10^-8^ threshold, AFR to EUR was 9 out of 25 vQTLs, and CSA to EUR was 8 out of 11 vQTLs, all with concordant effects. Comparisons between AFR and CSA were underpowered but replication was still observed at P_VE_ < 0.05, with AFR to CSA giving 9 out of 17 vQTLs and CSA to AFR 8 out of 21 vQTLs (**Figure S5**). Perhaps unsurprisingly, our findings highlight the variability of transferability and replicability of vQTL effect sizes and the importance of ancestry-stratified analyses for GEI interactions of vQTLs.

**Figure 2:**
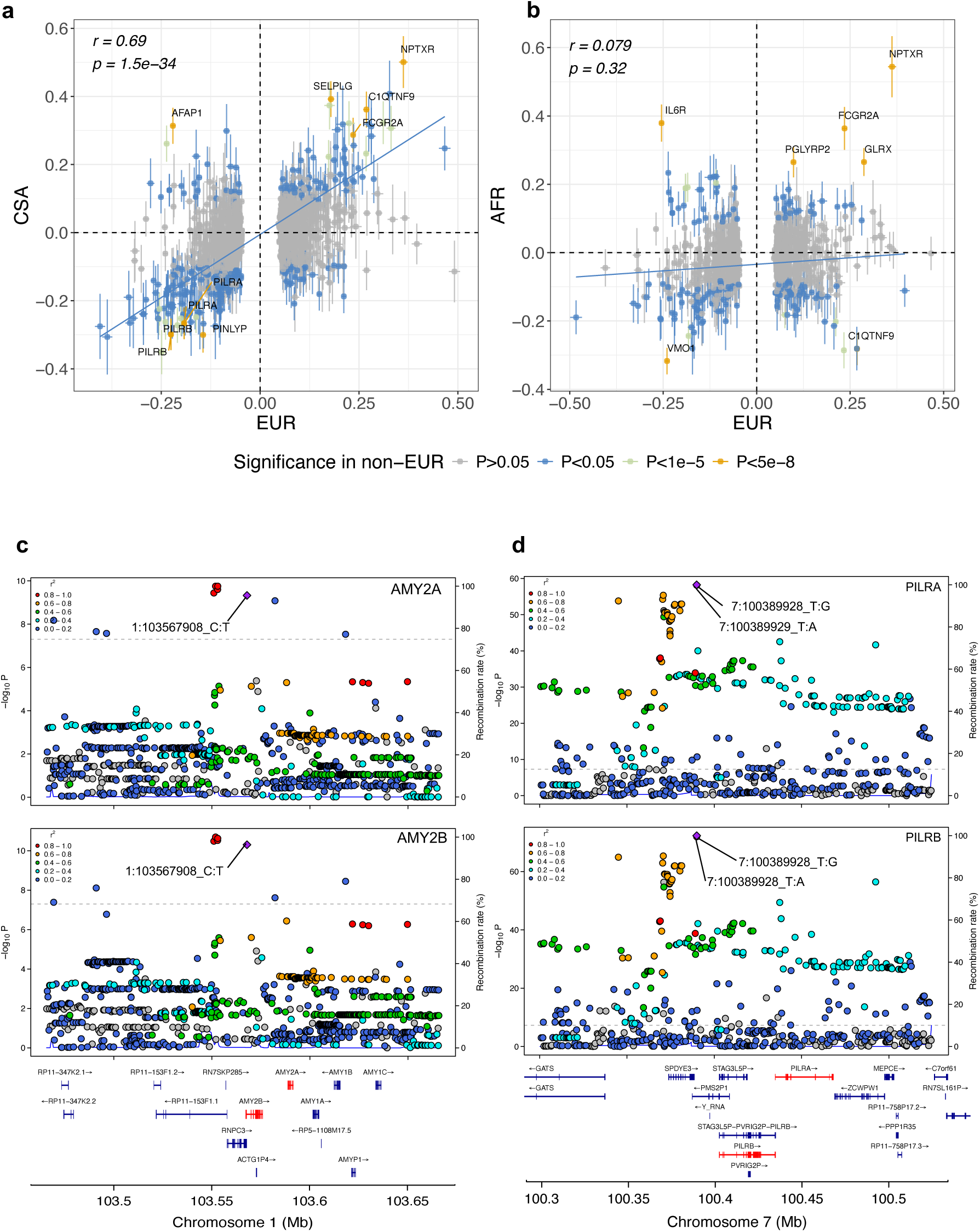
Comparison of vQTL associations across ancestries. Panels (**a-b**) show cross-ancestry correlation analysis of vQTL effect sizes to evaluate consistency in direction and magnitude. For each comparison, signals identified in the EUR were protein-matched and mapped to a non-EUR ancestry and restricted to those with MAF > 5% in the target population. The blue regression line and Pearson’s correlation estimate in the top left corner, are based on vQTLs that replicated at the nominal significance threshold (P_VE_ < 0.05) with concordant effects in the respective non-EUR population (points coloured blue). Only vQTLs that replicated at P_VE_ <5×10^-8^ in the non-EUR are annotated with their respective proteins (points coloured orange). Panels (**c-d**) LocusZoom plots showing evidence of genetic colocalization of vQTLs in the EUR, between AMY2A and AMY2B and PILRA and PILRB respectively.

Further, our findings supported the potential power of vQTL analyses in non-European ancestries. For proteins with independent vQTLs identified in both EUR and at least one non-EUR population (**Figure 1b**), we compared the strength of association signals by plotting the corresponding -log10 p-values (P_VE_). Despite the much larger sample size in EUR, for seven proteins we observed a stronger vQTL association in AFR (APOL1, CD86, DPEP, GCACT, ITIH3, SCLY and SIGLEC9) owing to greater estimated effect size or allele frequency (**Figure S6**). This was exemplified by the *APOL1* locus, which exhibited a highly significant independent vQTL in AFR (22:36660975_A:G, P_VE_ = 7.7 x 10^-^²⁶, Estimate_VE_ = −0.428416, S.E_VE_ = 0.04), compared to EUR (22:36652889_T:G, P_VE_ = 7.9 x 10^-^¹¹, Estimate_VE_ = −0.0472081, S.E_VE_ = 0.01), consistent with the well-known ancestry-specific effects at this locus^20^ despite stark differences in statistical power.

We also queried vQTLs from EUR and assessed their overlap with fine-mapped, ancestry and protein-matched *cis*-pQTLs. Specifically, we selected a parsimonious set of putatively causal *cis*-pQTLs with posterior inclusion probabilities (PIPs) > 0.9 from Sun et al.^3^ for 586 proteins where vQTLs were also detected. Although the majority of vQTLs (∼92% in EUR) exhibited both mean and variance effects, few vQTLs overlapped with putatively causal *cis*-pQTLs directly (i.e. same variant). Among the 2,001 vQTLs identified across 771 proteins, only 45 vQTLs (for 44 proteins) overlapped with fine-mapped *cis*-pQTLs that had PIP > 0.9 for mean effects, and this included just one variant with vQTL_only_ effects (4:6623342_T:C, MAN2B2; P_VE_ = 2.3×10^-19^). In addition, for proteins where a vQTL was detected that did not correspond to the same fine-mapped *cis*-pQTL variant, the average pairwise LD between the two sets of variants was low, with a mean *r*^2^ of 0.18 (**Figure S7**). Overall, only 3% of EUR vQTLs have high confidence overlap with putatively causal *cis*-pQTLs for the same proteins, indicating that most vQTLs influence protein variability independently of mean expression levels (**Table S4**).

Within each ancestry group, we also performed colocalization analyses to pinpoint evidence of shared vQTLs for different proteins. We used two complementary approaches: (i) first, we screened for colocalization among protein pairs that shared the same vQTL (based on duplicated vQTL signals), and (ii) we then examined proteins with vQTLs on the same chromosome that had overlapping TSS, defining region boundaries using the minimum gene start and maximum gene end positions to capture both associations and tested for evidence of colocalization (**Table S5**). Our results indicated only two of 19 pairs of proteins tested in EUR showed evidence of colocalization for vQTLs (H4 PP > 0.8). Interestingly, the two protein pairs showed evidence of colocalization for both mean and variance effects, separately. For AMY2A and AMY2B, we observed an H4 PP of 0.99 for variance effects, with the vQTL (1:103567908_C:T) also showing a SNP-level H4 PP of 0.99 (posterior probabilities for each SNP to be causal conditional on H4 being true). For mean effects, the shared vQTL signal (1:103602451_C:T) also had H4 PP and SNP H4 PP = 0.99. Similarly, for PILRA and PILRB, we observed H4 PP = 0.99 for variance effects due to two genetic variants: 7:100389928_T:G (SNP H4 PP = 0.25) and 7:100389929_T:A (SNP H4 PP = 0.75). For mean effects, the shared signal was 7:100366917_A:G, with both H4 PP and SNP H4 PP = 0.99 (**Figure 2c-d**, **Table S5**). In both cases, the variants identified through colocalization were however, not the same as the lead independent vQTLs, and were not in high LD with them (*r*^2^ < 0.2). We also identified one protein pair that showed evidence of colocalization in the mean effects and not in the variance effects (CGREF and KHK, H4 PP = 0.99, **Table S5**); however, no instances were identified where colocalization occurred exclusively in the vQTLs and not for the corresponding mean effects. Finally, we investigated instances of colocalization between mean and variance effects among proteins showing vQTL associations across ancestries (H4 PP > 0.8). Specifically, there were 134 in EUR, 10 in AFR, and 4 in CSA, with one overlap in both AFR and EUR (CAPG; AFR H4 PP: 0.99; EUR H4 PP: 0.99), and CSA and AFR (TLR3; AFR H4 PP: 0.99; CSA H4 PP: 0.86) (**Table S6**).

### Cross-ancestry gene-by-environment interaction analysis

We performed targeted GEI analysis focusing on the 30 proteins that exhibited vQTL overlap across the three ancestry groups (**Figure 1b-c**). We incorporated five common demographic and behavioural factors as exposures from UKB, namely: age, alcohol consumption, body mass index (BMI), sex and smoking status. To identify significant GEI, we applied a Bonferroni correction incorporating the 228 vQTLs tested, five exposures and three ancestries (*p* < 1.4×10^-5^) (**Methods**). After correcting for multiple testing across all vQTL-exposure combinations and ancestry groups, we assessed the interaction terms and identified three proteins with vQTLs (all with mean and variance effects) that passed the significance threshold in EUR. Notably, for DKKL1, we detected significant interaction effects across three exposures: age, sex, and BMI for the same variant (19:49788205_C:T; *p* = 4.0 x 10^-10^; 9.2 x 10^-13^, and 9.1 x 10^-14^, respectively). Additionally, PDGFRB exhibited significant interaction effects with age (5:149503670_A:G; *p* = 1.7 x 10^-6^), while a vQTL for TREH exhibited significant interactions effects with both age and BMI (11:118486067_C:T; *p* = 1.0 x 10^-6^ and 3.6 x 10^-14^) (**Figure S8** and **Table S7**). No significant interaction effects were detected in AFR and CSA, likely due to the lack of power compared to EUR.

### Proteome-wide Multivariable Mendelian randomization

Using vQTLs from EUR, we investigated whether genetically predicted variance of plasma protein levels causally influenced risk of coronary artery disease (CAD) or Type 2 diabetes (T2D). Our approach assessed whether variance effects were independently associated with CAD and T2D risk, conditional on genetically predicted mean levels (mean effects). To do this, we applied a multivariable Mendelian randomization framework (MVMR), jointly modelling mean and variance effects as two separate exposures against each disease outcome. GWAS summary statistics for CAD and T2D were obtained from participants of European ancestry in Aragam et al.^21^ (181,522 cases; 984,168 controls), and Suzuki et al.^22^ (242,283 cases; 1,569,734 controls), respectively.

We first conducted colocalization analysis to determine phenotypic heterogeneity, defined here as the differences in the effects of distinct genetic variants at a single locus on the two aforementioned exposures^17^. Using a colocalization analysis^23^ threshold of H3 / (H3 + H4) > 0.95, we screened all proteins with vQTLs and identified 530 proteins showing distinct, non-overlapping associations for mean and variance effects in EUR. We then applied MVMR using the Robust Principal Component analysis based Generalized Method of Moments (PC-GMM) that is able to disentangle the correlated yet distinct effects of trait mean and variance (**Methods**)^24^.For each outcome and protein locus without evidence of a shared causal variant for mean and variance effects, we selected the number of principal components that explained 99.9% of the weighted genetic variation at the locus^17^. Results from the analysis are presented in **Figure 3**, **Table 1**; **Figure S9**, **Tables S8-S9**. Effect estimates were expressed in log odds ratios per 1 standard deviation in either the genetically predicted levels (Effect_ME_) or variance (Effect_VE_) of the protein.

**Figure 3:**
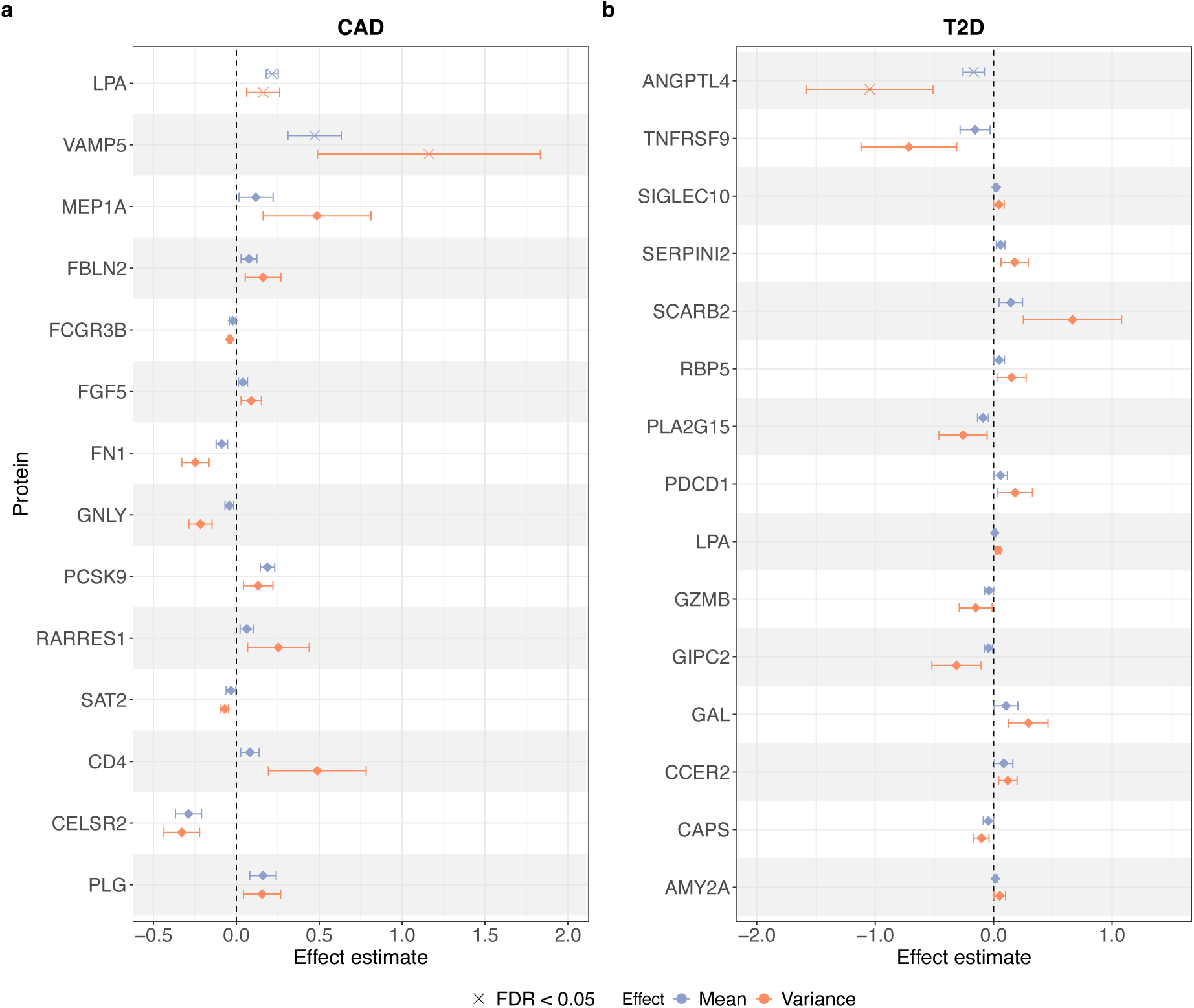
Proteins implicated by MVMR in CAD and T2D risk through mean and variance effects. The forest plots illustrate proteins with consistent effect directions for both mean and variance effects in CAD (a) and T2D (b), respectively. Significant results presented, show that genetically predicted variance is independently associated with the outcomes conditional on genetically predicted mean levels. Effect estimates are expressed in log odds ratios per 1 standard deviation change in the genetically predicted mean and variance of proteins. For the 29 proteins shown (14 for CAD, 15 for T2D), both protein variances and mean levels are significant at a nominal (P < 0.05) threshold. Lp(a) and VAMP5 for CAD, and ANGPTL4 for T2D, are denoted with an (x) and plotted at the top of panels (**a**) and (**b**) respectively, as showing significant associations for both mean and variance effects after Benjamini-Hochberg FDR correction (FDR < 0.05).

**Table 1:**
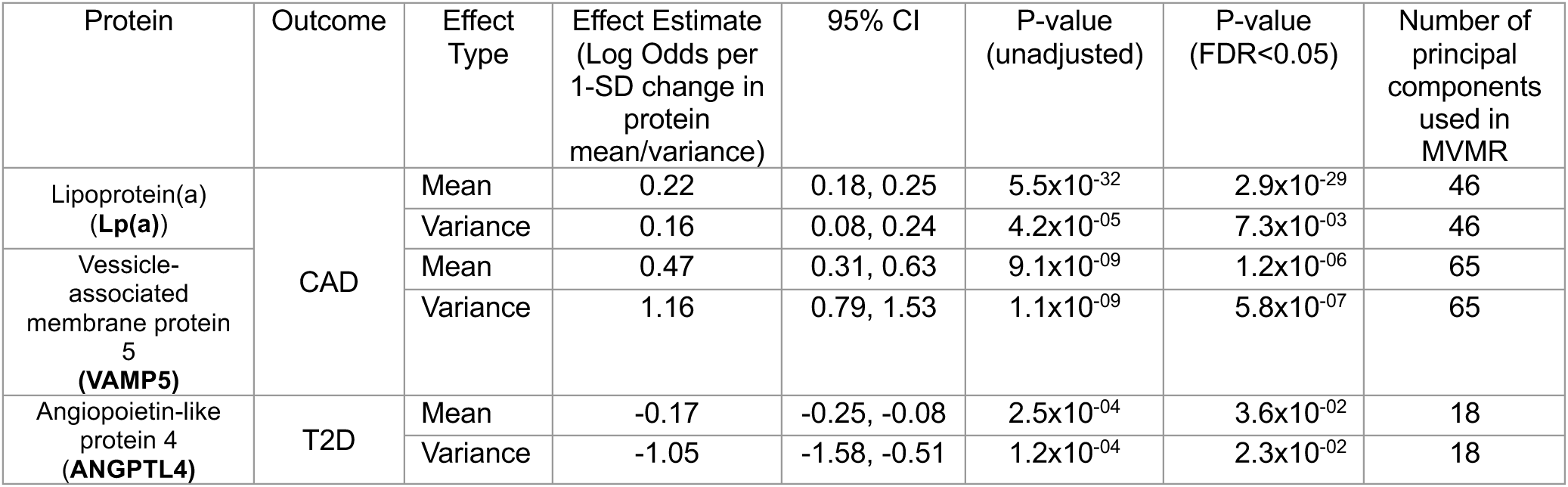
Genetically predicted multivariable mean and variance effects of proteins on CAD and T2D.

Interestingly, when filtering for nominal p-value threshold (P < 0.05) for both mean and variance effects, 29 of the 30 proteins detected exhibited consistent directions of effect, with the exception being CEACAM16 in T2D (Effect_VE_ = −0.10, P_VE_ = 4.1x 10^-3^, Effect_ME_ = 0.02, P_ME_ = 4.7 x 10^-2^) (**Figure S9a-b, Tables S8-S9**). Fourteen proteins showed evidence of causal association with CAD and another 15 with T2D, with Lp(a) showing positive effects of both mean and variance on both CAD and T2D (**Figure 3, Table1, Tables S8-S9**). Beyond Lp(a), variance effects of multiple drug targets for CAD were found, including PCSK9, fibronectin (FN1) and plasminogen (PLG) (**Figure 3, Table S8**). Three proteins passed Benjamini-Hochberg FDR <0.05 for both mean and variance effects, all of which are key proteins previously implicated in disease risk: Vesicle Associated Membrane Protein 5^25,26^ (VAMP5) (P_VE_ = 5.8×10^-7^, P_ME_ = 1.2 x 10^-6^) and Lp(a)^27,28^ (P_VE_ = 7.2 x 10^-3^, P_ME_ = 3 x 10^-29^) for CAD as well as Angiopoietin-like 4^29,30^ (ANGPTL4) for T2D (P_VE_ = 2.3 x 10^-2^, P_ME_ = 3.6 x 10^-2^) (**Figure 3**, **Table 1**; **Table S8-S9**). Similar to Lp(a), VAMP5 was positively associated with CAD risk (Effect_VE_ 1.16, 95% CI 0.79, 1.53; Effect_ME_ 0.47, 95% CI 0.31, 0.63). On the other hand, variance and mean effects of ANGPTL4 were inversely associated with T2D risk (log odds ratio per 1 standard deviation increase in both genetically predicted plasma ANGPTL4 variance and mean levels; Effect_VE_ −1.05, 95% CI −1.58, −0.51; Effect_ME_ −0.17, 95% CI −0.25, - 0.08) (**Figure 3**, **Table 1**).

For the VAMP5 and ANGPTL4 associations (with CAD and T2D, respectively), the effect estimates for variance were stronger and more statistically significant than the mean effects, indicating protein variability is a particularly important yet underexplored aspect of these putative therapeutic targets.

One association exhibited nominally significant protein variance effects on disease in the absence of significant mean effects (**Figure S9**, **Table S9**). Variance in levels of circulating Junction Adhesion Molecule 3 protein (JAM3) were associated with T2D risk (P_VE_ = 4.4 x 10^-6^, P_ME_ = 5.3 x 10^-1^; log odds ratio per 1 standard deviation increases in genetically predicted plasma JAM3 variance levels; Effect_VE_ 0.36, 95% CI 0.20, 0.51) but the genetically predicted mean levels were not (Effect_ME_ 0.019, 95% CI −0.05, 0.09).

## Discussion

Our multi-ancestry vQTL analysis of the human proteome makes two major advances. First, we identified protein vQTLs beyond European populations to Central/South Asian and African ancestry groups, substantially increasing the number of known protein vQTLs overall. Second, we showed the independent putatively causal effects of protein levels and protein variances on two major common disease, type 2 diabetes and coronary artery disease. We also highlight several key findings: enrichment of vQTLs on chromosome X relative to the autosomes; variable transferability of vQTL effect sizes across ancestries; and limited horizontal pleiotropy among *cis-*vQTLs, with only two protein-pairs in the EUR, showing evidence of colocalization for both variance and mean effects within the same genomic regions. Additionally, we find evidence of colocalization between mean and variance effects for proteins with vQTL associations across all ancestries, one showing overlap in AFR and CSA, and another in EUR and AFR. We also identified the largely distinct genetic architectures of protein means and variances by finding limited overlap between vQTLs and fine-mapped variants driving mean effect associations in proteins, despite the majority of vQTLs exhibiting mean effects.

A previous landmark study, Hillary et al^11^, assessed autosomal *cis*-vQTLs in EUR ancestry participants of UK Biobank and reported 421 *cis*-vQTLs for 423 proteins from the initial 1,472 Olink protein set, approximately one *cis*-vQTLs per protein. By way of comparison, this current study considers the 2,923 Olink protein set and identifies 2,162 *cis*-vQTLs for 781 proteins (approximately 2.7 *cis*-vQTLs per protein). We replicated 96.9% (408/421) of the near-independent *cis*-vQTLs reported by Hillary et al for their respective proteins in our EUR analysis. Of these, 56% (236) were replicated directly for the same variants and proteins, while the remainder were captured via their pairwise LD proxies (0.99 > *r^2^* > 0.1) (**Table S10**).

Our findings suggests that changes in protein variance, alongside changing protein levels, may have therapeutic potential. From our MVMR analysis, ANGPTL4 emerged as the most promising candidate associated with T2D. ANGPTL4 is an endogenous inhibitor of lipoprotein lipase (LPL), playing a key role in lipid regulation, coronary atherosclerosis risk, and nutrient partitioning^29,30^. Consistent with our findings, genetic inactivation of ANGPTL4 has also been linked to improved glucose regulation and reduced risk of T2D^31^. The Open Targets Genomics platform shows an association score of 0.87 for ANGPTL4 with T2D, indicating strong support from multiple lines of evidence for this association^32^. While some MR studies have implicated ANGPTL4 and T2D association based on trait mean levels^33,34^, this is the first study implicating ANGPTL4 variance as an independent risk factor for T2D. VAMP5 and Lp(a) have previously been implicated in CAD based on mean effects, with cumulative evidence drawn from multiple experimental and *in silico* studies^25–28^. The Open Targets Genomics platform association scores with CAD were 0.58 and 0.92 for VAMP5 and Lp(a), respectively^32^. For both proteins, our MVMR results suggest that increasing protein variance in turn increases risk of CAD independent of the protein levels.

More proteomic data in globally diversity populations is urgently needed. We found proteins with vQTLs that were shared across ancestries, and, in multiple cases, stronger signals were observed in non-EUR ancestries, despite EUR being better powered for discovery. The *APOL1* locus, which has been strongly associated with increased risk of kidney disease particularly among individuals of African descent^35–38^, is a key example. Risk variants in *APOL1* have been linked to higher rates of end-stage renal disease and faster progression of chronic kidney disease in individuals of African ancestries compared to those of European ancestries^35^. Our ancestry-enriched association for APOL1 is noteworthy as variability in APOL1 levels could contribute to the observed disparities in renal disease outcomes and may reflect underlying ancestry-specific genetic architecture. The APOL1 signal in AFR displayed variance effects alone, as it did not have even nominally significant mean effects. In EUR, the signal identified had both mean and variance effects but was in low pairwise LD with the AFR signal. In our data, the known CKD-associated allele G1 showed a strong association with APOL1 in AFR (22:36661906; P_VE_ = 8.7 x 10^-19^, P_ME_ = 1.02 x 10^-3^); however, the G2 allele, a six nucleotide deletion^36^, could not be tested due to quality filters. Further investigation of the multiple vQTL, including G1, effects at the *APOL1* locus may dissect the role trait variance plays in shaping APOL1-mediated kidney disease in African populations.

Our investigations have several limitations. Most proteomic data is available only in cohorts that are largely European, limiting both diversity and replication opportunities for non-European populations. This invariably affected the number of signals detected in the non-EUR ancestries and their cross-ancestry replication rates. Our MVMR analyses in non-European ancestries also failed to identify significant associations, likely reflecting limited statistical power. Additionally, even though we observed proportional enrichment of vQTLs on chromosome X compared to the autosomes, further studies utilising RNA-seq data will be needed to explore the extent to which vQTLs and chromosome X-inactivation signals are shared or distinct.

In conclusion, we present a comprehensive characterization of protein vQTLs across diverse ancestries, underscoring the role that trait variance plays in shaping the genetic architecture of the human proteome. We further uncover the distinct and complementary effects of protein levels and variances on disease outcomes, which may have implications for target prioritisation and therapeutic development.

## Methods

### Study cohort

We studied participants from the UK Biobank study (UKB), a long-term prospective population-based cohort of approximately 500,000 adults aged between 40 and 69 years, recruited between 2006 and 2010 across 22 recruitment centres in the UK^39^. In this study, a subset of the UKB sample was used as defined by the UK Biobank Pharma Proteomics Project (UKB-PPP) consortium. Briefly, the sample comprised of 54,219 participants broken down as follows: (1) a randomized subset of 46,595 individuals; (2) 6,356 individuals selected by the UKB-PPP consortium members (‘consortium selected’), in which the initial proteomic profiling was done from the baseline assessment and (3) 1,268 individuals who participated in a COVID-19 repeat imaging study. Detailed descriptions of the sample selection and handling procedures have been described elsewhere^3^. The current analysis was approved under UK Biobank Project application number 7439.

Genotyping details for the UK Biobank cohort have been reported previously^39^ and were performed by the UKB. Briefly, two custom genotyping arrays were utilised with 49,950 participants typed using the UK BiLEVE Axiom Array and 438,427 participants typed using the UK Biobank Axiom Array. The two arrays shared 95% coverage resulting in 805,426 genotyped markers on 488,377 participants. Imputation was carried out centrally by the UKB, primarily using the HRC+UK10K haplotype resource and IMPUTE4^40,41^. Analysis in this study was conducted with version 3 of the UKB imputed data with 487,442 participant samples imputed and 93,095,623 autosomal SNPs available for analysis following UKB centrally performed QC filters. Imputed genotypes were based on GRCh37 coordinates.

### Genetic similarity analysis

We stratified the UKB participants (*N* = 52,706) into sub-populations based on genetic similarity using KING^42^. Briefly, genotyped UKB participant samples were projected onto the top 10 genetic principal components (PCs) computed from the 1000 Genomes Project’s phase 3’s (1KGP) five super-populations: African (AFR); Admixed American (AMR); East-Asian (EAS); European (EUR) and Central/South Asian (CSA). Based on these projections, KING utilizes a support vector-machine-based method to infer the most likely ancestral group for each participant. We refer to UK Biobank ancestry assignments using the 1KGP superpopulation labels to describe genetic similarity in accordance with recent reports that explored population descriptors in much greater detail^18^, e.g. 1KGP-AFR-like for participants that cluster closer to AFR superpopulation clusters. For brevity, we shorten these labels with three-letter population codes (AFR, EUR and CSA) throughout this manuscript.

We first applied standard QC filters to genotyped variants (MAF ≥ 0.01, HWE p-value ≥ 1e-6, missingness ≤ 0.1, genotyping rate ≥ 0.1) and generated an LD-pruned set using PLINK1.9^43^ (--indep-pairwise 50 5 0.2), retaining SNPs with pairwise correlation *r*^2^ < 0.2 within a 50-SNP window, sliding forward in increments of 5 SNPs at a time. Variants in long-range LD regions were also excluded (chr6: 25-33.5 Mb, chr8: 8-12 Mb, chr17: 40.4-42.4 Mb). Parameters used in KING were the “--pca” and “--projection” flags on the merged UKB-1KGP superpopulations dataset.

UKB participants were assigned to the EUR ancestry group if their probability of belonging to the corresponding superpopulation group (Pr) was ≥ 0.99; for AFR and CSA, a slightly relaxed threshold was used for ancestry assignment (Pr ≥ 0.95). The relaxed threshold used in the non-EUR ancestry groups was due to comparatively smaller sample sizes and to allow us to capture more participants within each non-EUR ancestry group. We prioritised these three subpopulations due to their relatively larger sample sizes, ensuring sufficient power for discovery (**Figure S2**). Subsequently, we excluded related participants within each subpopulation by using PLINK2.0^43^ with the “--king-cutoff 0.0884” argument to remove one individual from each pair of first- or second-degree relatives. We then recalculated PCs and retained participants who fell within ±4 standard deviations of the top 4 PCs in each ancestry, separately. The final pruned sample lists (AFR, *N* = 1,336; EUR, *N* = 45,486; CSA, *N* = 933) was used to compute twenty ancestry-specific PCs and for the downstream stratified analysis.

### Proteomic quality control and phenotype modelling

Proteomic profiling was conducted centrally by the UKB-PPP on EDTA-plasma samples from 54,306 participants using the Olink Explore 3072 platform. Protein quantification was achieved through the Olink Proximity Extension Assay (PEA) combined with next-generation sequencing. Sample selection for the proteomic assay by the UKB-PPP was based on several factors, as described in detail elsewhere^3^. The Olink assay technology and analytical methods are also detailed in a separate publication^3^. In brief, the relative abundance of 2,923 proteins was quantified using antibodies distributed across four 384-plex panels: cardiometabolic, inflammation, neurology and oncology. Measurements were expressed as normalised protein expression (NPX) values that were log-base-2 transformed. Protein values below the limit of detection (LOD) were replaced with the LOD divided by the square root of 2 and each protein was rescaled to have a mean of 0 and a standard deviation (SD) of 1.

We modelled our protein phenotypes by regressing out the effects of the following covariates: age, age^2^, sex, age*sex, age^2^*sex, batch (b0 - b7), UKB assessment center, UKB genotyping array, time between blood sampling and measurement and the first 20 genetic principal components (PCs). Subsequently we performed inverse normal transformation of the residuals, which we then used as phenotypes in our analysis. Previous publications, such as Wang et al.^9^ have suggested that applying inverse normal transformation (INT) may reduce power for discovery in complex traits. However consistent findings from Hillary et al.^11^, where INT was applied to protein levels in a similar context suggest that effect estimates before and after transformation are highly correlated (*r* = 0.9). Subsequently, we conducted a sensitivity analysis which yielded similar results (**Figure S10**, **Tables S10-S11**).

### *C*is-vQTL mapping

We ran quality control (QC) filters on imputed dosages (MAF ≥ 0.01, HWE p-value ≥ 1 x 10^−6^, imputation info score R^2^ ≥ 0.3) for bi-allelic autosomal markers using PLINK1.9^43^ for each ancestry. Using OSCA^44^, we conducted two-sided, median-based vQTL mapping^9^. The test statistic of Levene’s test is:

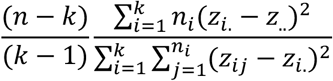

where *n* is the total sample size, *k* is the number of groups (k = 3 in vQTL analysis), *n_i_* is the sample size of the *i*th group (one of three genotypes), *z_i,j_* is the absolute difference between the phenotype value in sample *j* from genotype and the median value in genotype *i*, *z_i_*_._ is the average *z* value in genotype *i*, and *z*_.._ is the average *z* value across all samples. The OSCA implemented Levene’s test also provides effect estimates and standard errors estimated using p-values and minor allele frequencies.

### Locus definition and variant annotation

To identify near-independent associations, we applied LD and distance-based clumping implemented in PLINK1.9^43^ with parameters--clump-r2 0.01, --clump-kb 1000, --clump-p1 5 x 10^−8^ and --clump-p2 0.05, clumping variants ±1 Mb around significant vQTLs. We set a high confidence p-value threshold (P_VE_ < 5×10^-8^) for variants with MAF > 5% across ancestries. The MAF filter, was in accordance with the workflow of Wang et al^9^ in vQTL analyses to avoid the potential of false-positive associations due to the coincidence of a low-frequency variant with an outlier phenotype. We defined a *cis*-vQTL as a variant residing within 1Mb upstream or downstream (±1Mb) of the transcription site (TSS) of the corresponding protein-coding gene. We extracted the TSS information for all proteins from BioMart R package (accessed in October 2024) by mapping UniProt IDs to gene start, gene end and transcription start sites. For proteins with multiple transcripts, we selected the canonical 5’ TSS. To account for long-range LD that may have been overlooked by this approach, we also calculated pairwise LD between all vQTLs for proteins with multiple associations using PLINK1.9^43^.

vQTLs were annotated using the Ensembl Variant Effect Predictor (GRCh37.p13)^45^. We conducted enrichment analysis of vQTLs across functional classes using ANNOVAR from FUMA’s SNP2GENE function with default parameters, accounting for the different sample sizes of each ancestry group and using ancestry-matched reference panels (UKB release2, 10K European for EUR; 1000 Genomes Phase 3 AFR for AFR and 1000 Genomes Phase 3 SAS for CSA). The nearest gene to the reported *cis*-vQTL was annotated using the python package GeneLocator (v.1.1.2), that returns a list of genes or overlapping genes in which the reported *cis*-vQTL is included, or the gene whose start or end is closest to the specified coordinates in the case a SNP does not fall within any genes.

### Chromosome X analysis

For each ancestry group, we included unrelated individuals inferred using KING^42^ with <10% missing genotypes and consistent sex based on chromosome X heterozygosity using PLINK1.9^43^. Pseudoautosomal regions (PAR1: 60,001-2,699,520 bp; PAR2: 154,931,044-155,260,560 bp) were also excluded. Variants with significantly different MAF or missingness rates between males and females were removed using Fisher’s Exact Test in R. Males (hemizygous) were recoded as 0/2, females as 0/1/2, and the sex-specific files were then merged for downstream chromosome X analysis.

### Gene-by-environment interaction tests

We conducted targeted gene-by-environment (GEI) analyses separately within each ancestry group, to assess whether vQTLs interacted with five common demographic and behavioural factors as exposures (age, sex, BMI, alcohol consumption, and smoking status) in influencing protein levels. We restricted our analyses to thirty proteins with vQTLs shared across all three ancestry groups, using a linear regression model implemented under the mixed linear model-based tool fastGWA^46^ with the linear regression flag “--fastGWA-lr flag” and specifying each exposure using the “--envir” flag. Since protein levels had already been adjusted for relevant covariates during protein modelling, we did not include any fixed effect covariates in our association model, in line with what is suggested in literature^47^. For their respective GEI models, sex and age were excluded as covariates during protein modelling. We defined the significance threshold using Bonferroni correction based on the number of tests performed: 228 vQTLs x 5 exposures x 3 ancestries, resulting in a corrected p-value threshold p = 1.4 x 10^-^⁵ to determine statistical significance. We then assessed the interaction terms of each vQTL-exposure pair, across ancestries for significance. The formula used for the interaction analysis was *Protein level ∼ SNP(vQTL) * exposure + SNP(vQTL) + exposure*

### Colocalization analysis

We identified shared vQTLs between all protein-pair combinations per ancestry, using the COLOC R-package^23^, under the assumption of single shared causal variant per protein-encoding locus. We performed colocalization analyses for proteins with overlapping gene start and end positions on the same chromosome (based on chromosome and bp position). For these protein pairs, we further tested for evidence of colocalization in the mean effects. Posterior probabilities (H4 PP) > 0.8 were used to classify significant associations. In instances where we identified evidence of colocalization, we reported the SNP-level H4 PP, which is the posterior probabilities for each SNP to be causal conditional on H4 being true (**Tables S5-S6**).

### Multivariable Mendelian Randomization

We conducted multivariable Mendelian Randomization (MVMR) using *cis*-genetic variants as instrumental variables to detect the direct independent effects of genetically predicted variance and mean levels of selected proteins as exposures, against CAD and T2D as outcomes^17^. MR estimates the total causal effect of the exposure on the outcome, whereas MVMR estimates the direct causal effect of each exposure on the outcome^48^. Briefly, within the MR framework, analysing too many variants at a single locus can produce spurious estimates, as correlations between variants can inflate the precision of causal effect estimates and obscure the independent contribution of each variant, while using too few instruments ignores most of the data and can also yield unstable estimates that are highly sensitive to the choice of variants in the model^49^. Dimension reduction techniques, such as principal component analysis, can help overcome some of these limitations by transforming correlated variants in a locus into a smaller set of uncorrelated components that capture most of the underlying genetic variation^17,50^.

To conduct the MVMR analysis we took the following steps: **(1)** For all proteins with significant vQTLs, we estimated the main genetic effects using PLINK2.0^43^, using the linear regression model flag (--glm), under an additive model. We did not adjust for additional covariates as covariate effects had already been regressed out during phenotype modelling of the proteins. **(2)** We conducted colocalization analysis using the “coloc_abf” function of COLOC ^23^ in R, using summary data for mean and variance effects for each protein with a significant vQTL as input, under the assumption of a single causal variant between traits. We applied default parameters and selected proteins where the posterior probability of distinct causal variants (H3) relative to shared variants (H4) was high (i.e., H3 / (H3 + H4) ≥ 0.95), suggesting non-overlapping genetic signals. We selected a total of 530 proteins which passed this test for downstream analysis in the EUR. **(3)** We retained variants with p <0.05 for both exposures and used the TwoSampleMR^51^ package in R and the “harmonize_data” function to harmonise the alleles and effects between the exposures (mean and variance effects of each protein) and the outcomes (CAD and T2D) in the *cis*-region. **(4)** We then generated linkage disequilibrium (LD) matrices for each protein gene region using LDstore2^52^. **(5)** For each protein, we used the “kriging_rss” function in SuSiE_RSS^53^ to detect possible allele switches and confirm alignment between alleles in the *cis*-region and the LD matrices generated. We did this for all 530 proteins. **(6)** For each protein and each outcome, we incorporated the corresponding LD matrix into the PC-GMM algorithm, implemented in the Mendelian Randomization R-package^24^, using effect estimates and standard errors from the variance and mean effects for protein levels, as well as the matching summary data for the retained variants from the respective outcomes as inputs. We used the robust version of PC-GMM and also set the PCA threshold to 0.999, which estimates the number of PCs that explain 99.9% of variation in the *cis*-region of the specific protein. We did this to account for the inclusion of multiple potentially correlated variants in the model, which could result in biased estimates, a limitation addressed by the PC-GMM framework^17,24^. **(7)** We set an additional filter post-hoc analysis, to filter proteins based on the nominal p-value threshold for both mean and variance effects (p < 0.05). Most proteins in either outcome showed consistency in directionality for effect estimates. Subsequently, we applied a Benjamini-Hochberg false discovery rate (FDR) correction and highlighted proteins with significant results at FDR < 0.05. To evaluate the robustness of this approach, we conducted sensitivity analyses by incrementally varying the PCA variance explained threshold, from 95% to 99% for three selected proteins that passed all tests listed in step 7 for CAD. While the number of principal components varied, significance of p-values remained consistent, supporting the stability of the model at the 99.9% threshold (**Table S12**). We also performed sensitivity analyses by reducing the window size around the TSS from 1MB to 500Kb for the proteins (LPA, VAMP5, ANGPTL4), which was largely insensitive to the smaller distances. The MVMR analysis used “mr_mvpcgmm” implemented in the MendelianRandomization R package^24^.

## Data Availability

All data produced in the present study are available upon reasonable request to the authors

## Acknowledgements

The authors are grateful to the UK Biobank for access to the data used in this study (Project #7439). This work was performed using resources provided by the Cambridge Service for Data Driven Discovery (CSD3) operated by the University of Cambridge Research Computing Service (www.csd3.cam.ac.uk), provided by Dell EMC and Intel using Tier-2 funding from the Engineering and Physical Sciences Research Council (capital grant EP/P020259/1), and DiRAC funding from the Science and Technology Facilities Council (www.dirac.ac.uk). This work was supported by core funding from the British Heart Foundation (RG/F/23/110103), NIHR Cambridge Biomedical Research Centre (NIHR203312) [*], BHF Chair Award (CH/12/2/29428), Cambridge BHF Centre of Research Excellence (RE/24/130011), and by Health Data Research UK, which is funded by the UK Medical Research Council, Engineering and Physical Sciences Research Council, Economic and Social Research Council, Department of Health and Social Care (England), Chief Scientist Office of the Scottish Government Health and Social Care Directorates, Health and Social Care Research and Development Division (Welsh Government), Public Health Agency (Northern Ireland), British Heart Foundation and the Wellcome trust. S.C.R is funded by the BHF Cambridge Centre for Research Excellence RE/24/130011. X.J. was funded by a Wellcome Trust Fellowship [227566/Z/23/Z). S.A.L. was supported by a Canadian Institutes of Health Research postdoctoral fellowship (MFE-171279). Y.X. and M.I. were supported by the UK Economic and Social Research Council (ES/T013192/1). The funders had no role in study design, data collection and analysis, decision to publish, or preparation of the manuscript. *The views expressed are those of the authors and not necessarily those of the NIHR or the Department of Health and Social Care.

## Declarations

M.I. is a trustee of the Public Health Genomics (PHG) Foundation, a member of the Scientific Advisory Board of Open Targets and has research collaborations with AstraZeneca and Nightingale Health which are unrelated to this study.

## Code Availability

The software used in this study can be assessed as follows: OSCA (v0.46.1): https://yanglab.westlake.edu.cn/software/osca/#vQTLAnalysis; Plink1.9 (v.1.90.b6.24): https://www.cog-genomics.org/plink/1.9/; Plink2 (v.2.0.a4): https://www.cog-genomics.org/plink/2.0/; COLOC (v.5.2.3): https://chr1swallace.github.io/coloc/; LDstore2 (v.2.0): http://www.christianbenner.com/#cmd_ld; SuSiE (v.0.14.21): https://stephenslab.github.io/susieR/index.html; FUMA (v.1.8.0): https://fuma.ctglab.nl/updates;GeneLocator(v.1.1.2):https://pypi.org/project/GeneLocator; TwoSampleMR (v.0.5.6): https://mrcieu.github.io/TwoSampleMR/;GCTA-fastGWA (v.1.95): https://yanglab.westlake.edu.cn/software/gcta/#fastGWA; KING (v.2.3.2): https://www.kingrelatedness.com/ancestry/; MendelianRandomization (v.0.10.0): https://cran.r-project.org/web/packages/MendelianRandomization/index.html. All custom scripts used in this study can be assessed through GitHub: https://github.com/Chiefeghan/vQTL_multi-ancestry

## Notes

### Funding Statement

The authors are grateful to the participants UK Biobank for access to the data used in this study (Project #7439). This work was performed using resources provided by the Cambridge Service for Data Driven Discovery (CSD3) operated by the University of Cambridge Research Computing Service (www.csd3.cam.ac.uk), provided by Dell EMC and Intel using Tier-2 funding from the Engineering and Physical Sciences Research Council (capital grant EP/P020259/1), and DiRAC funding from the Science and Technology Facilities Council (www.dirac.ac.uk). This work was supported by core funding from the British Heart Foundation (RG/F/23/110103), NIHR Cambridge Biomedical Research Centre (NIHR203312) [*], BHF Chair Award (CH/12/2/29428), Cambridge BHF Centre of Research Excellence (RE/24/130011), and by Health Data Research UK, which is funded by the UK Medical Research Council, Engineering and Physical Sciences Research Council, Economic and Social Research Council, Department of Health and Social Care (England), Chief Scientist Office of the Scottish Government Health and Social Care Directorates, Health and Social Care Research and Development Division (Welsh Government), Public Health Agency (Northern Ireland), British Heart Foundation and the Wellcome trust. S.C.R is funded by the BHF Cambridge Centre for Research Excellence RE/24/130011. X.J. was funded by a Wellcome Trust Fellowship [227566/Z/23/Z). S.A.L. was supported by a Canadian Institutes of Health Research postdoctoral fellowship (MFE-171279). Y.X. and M.I. were supported by the UK Economic and Social Research Council (ES/T013192/1). The funders had no role in study design, data collection and analysis, decision to publish, or preparation of the manuscript. *The views expressed are those of the authors and not necessarily those of the NIHR or the Department of Health and Social Care.

### Author Declarations

All data described are available through the UK Biobank subject to approval from the UK Biobank access committee. See https://www.ukbiobank.ac.uk/enable-your-research/apply-for-access for further details

