## Supplementary_Figures for "The genetic determinants of plasma protein variance across ancestries and effects on cardiometabolic disease risk"

This file contains:

Supplementary Figures 1-10

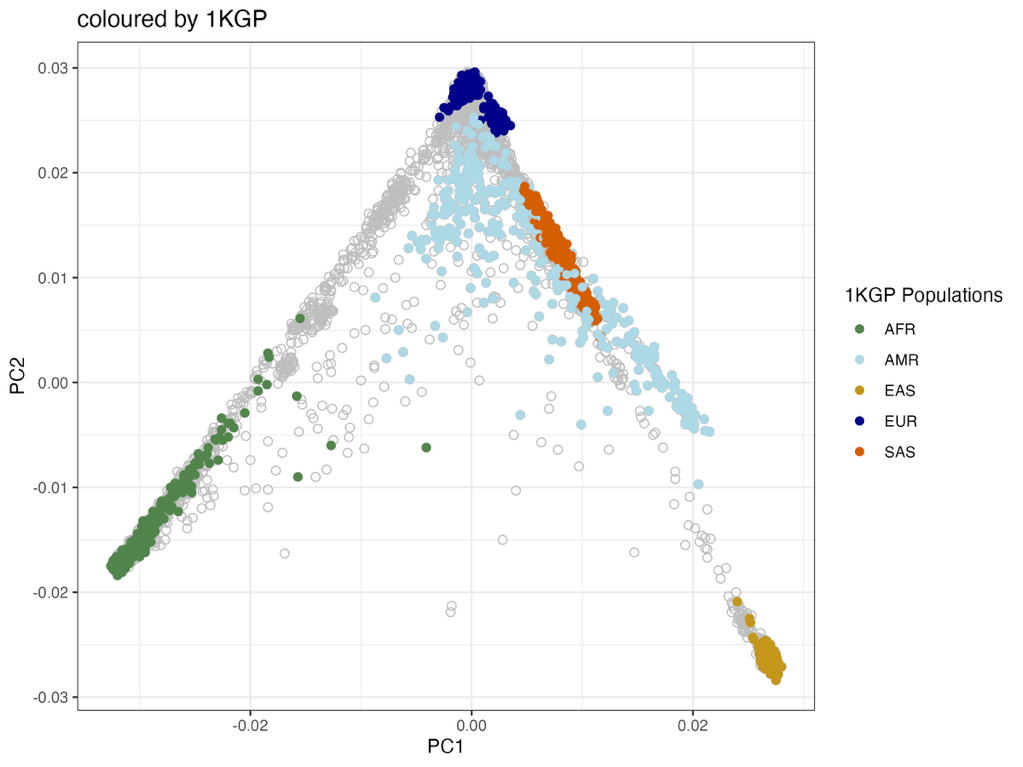

**b**

**a**

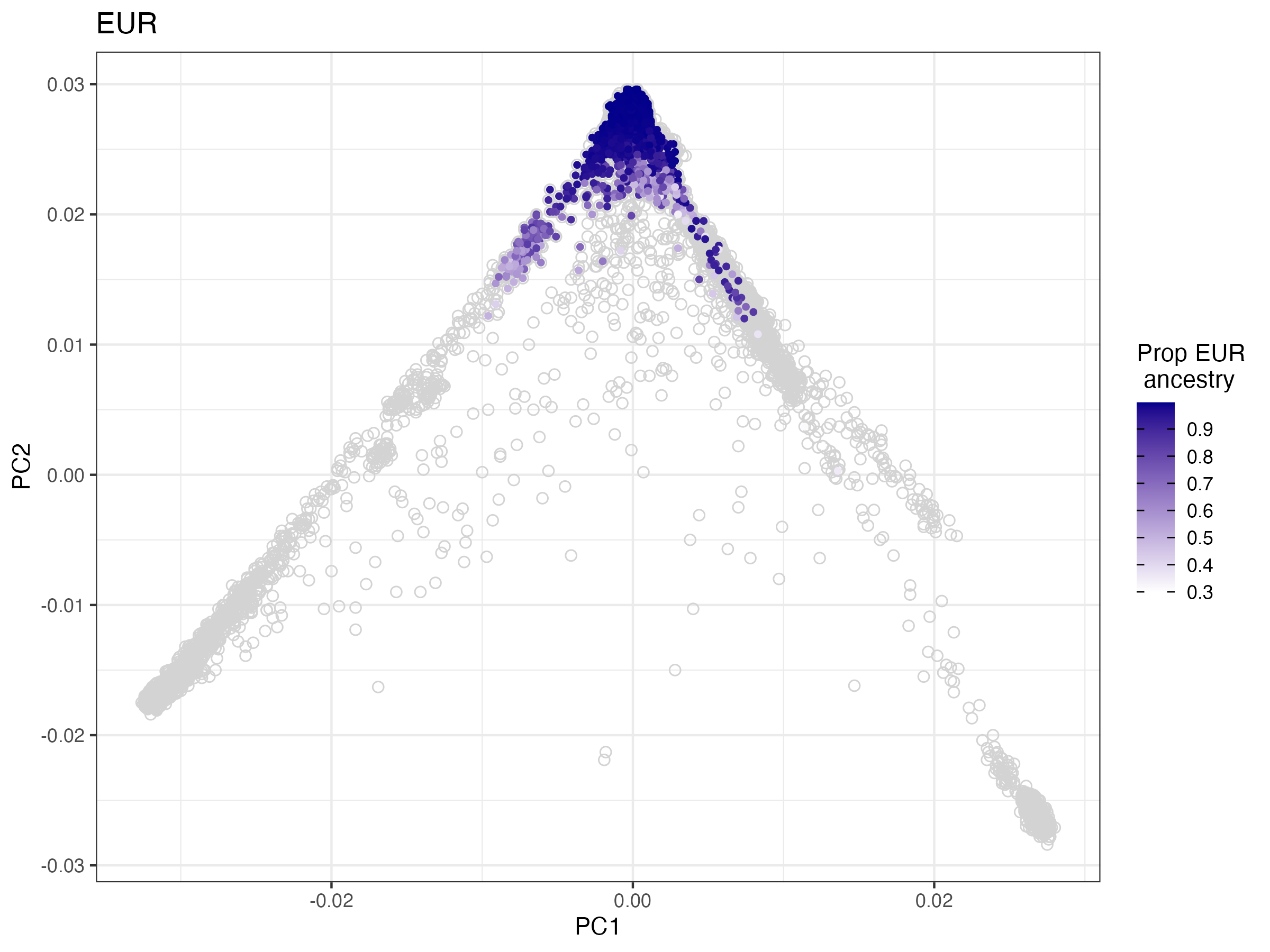

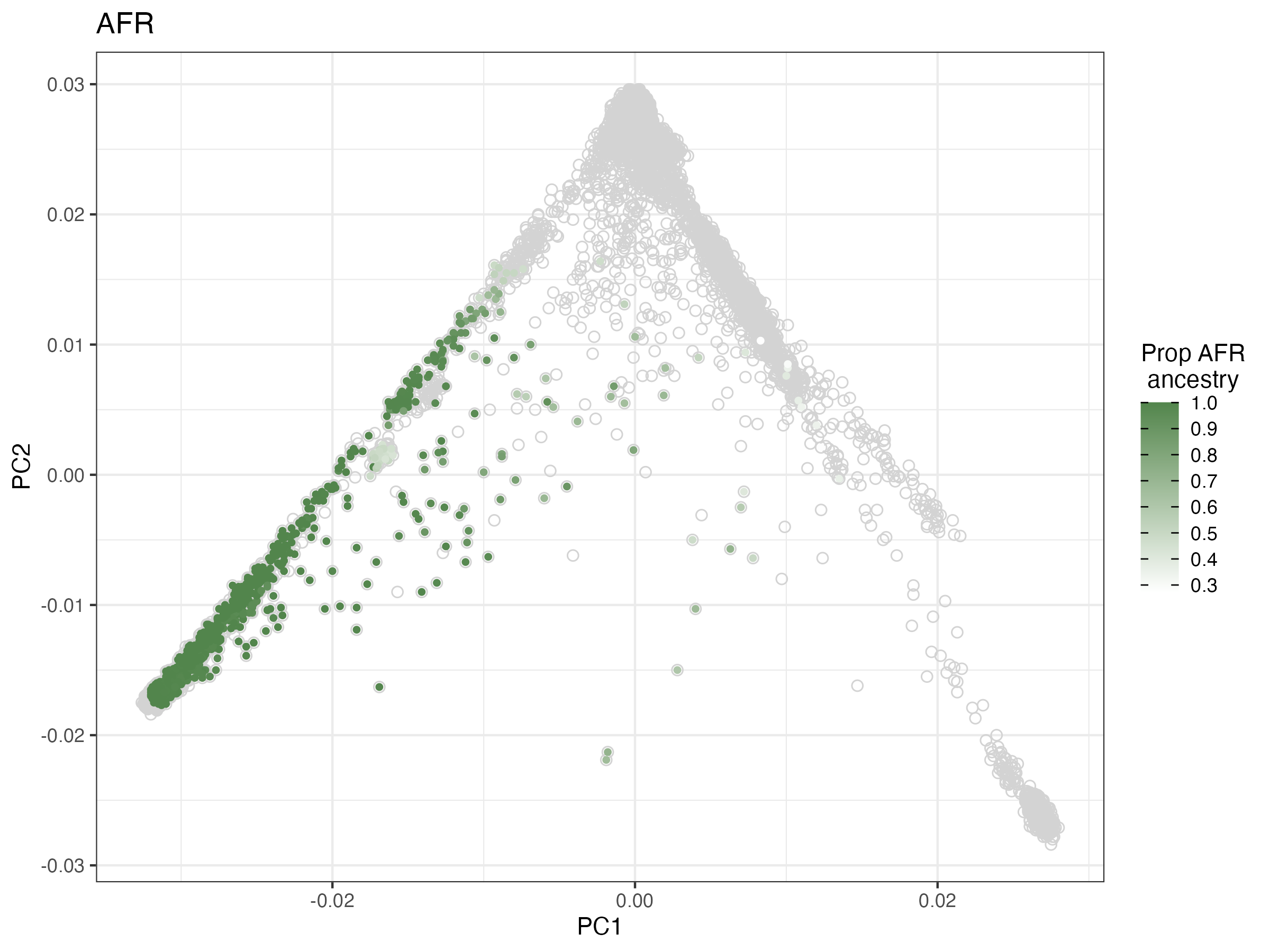

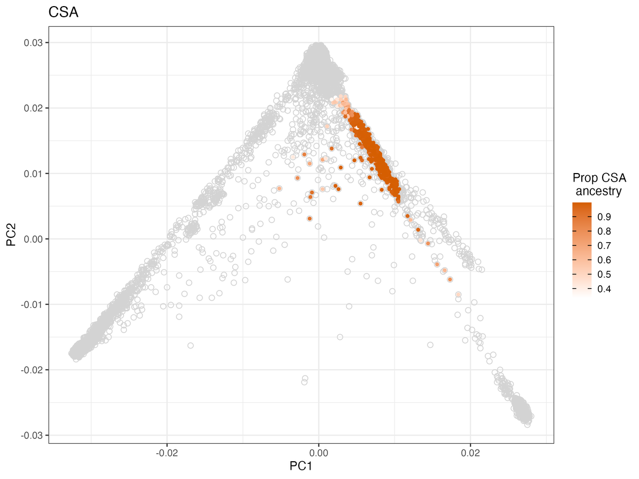

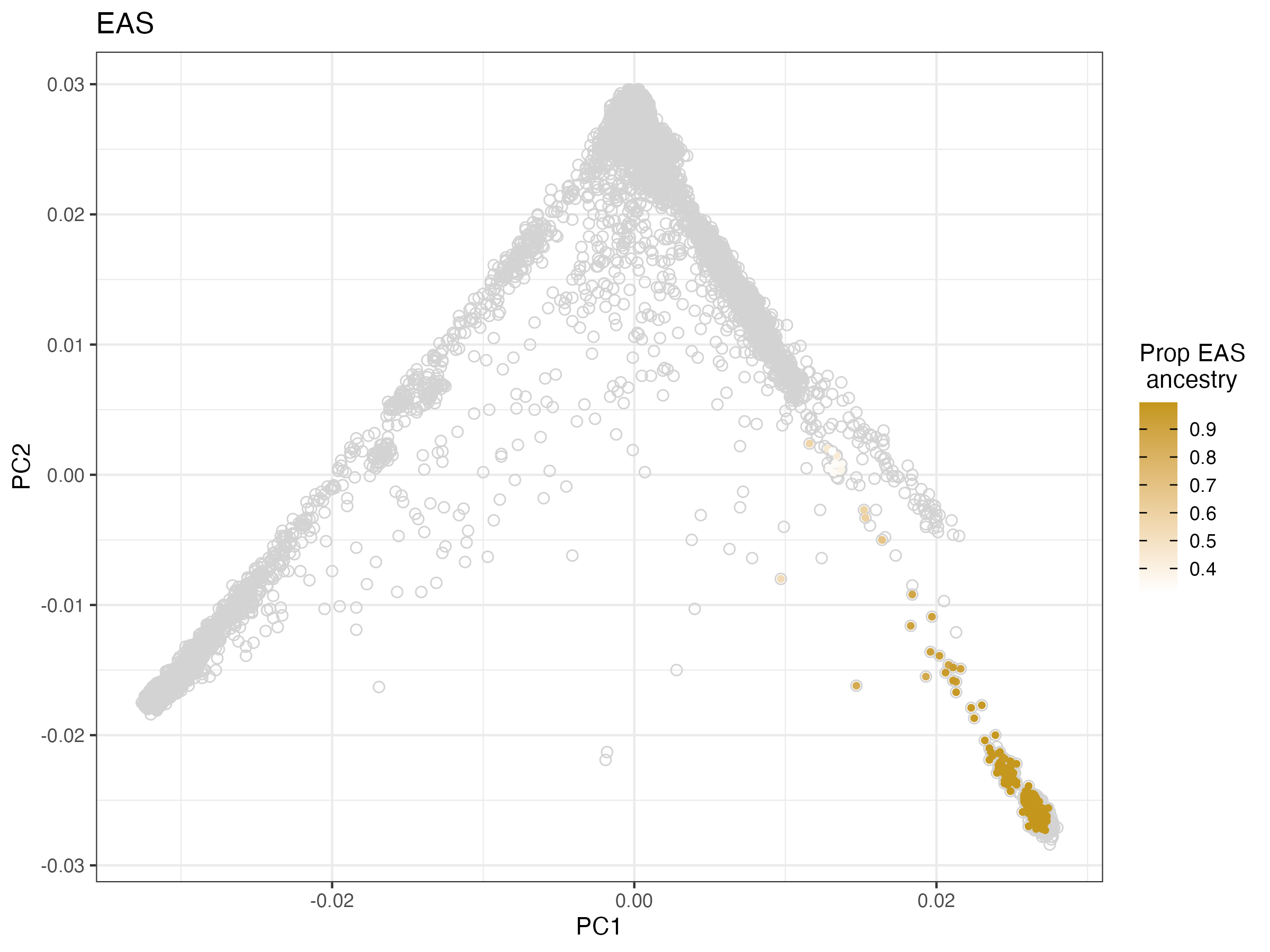

**e**

**d**

**c**

**Figure S1**. **Ancestry inference of UKB samples**: Panel (a): UKB samples were merged with 1000 Genomes Project Phase 3 (1KGP) reference data and projected onto the top 10 genetic principal component space. Samples are coloured by 1KGP super-populations: African (AFR), Admixed American (AMR), East Asian (EAS), European (EUR), and Central/South Asian (CSA); grey indicates samples that did not cluster closely with any reference group. Panels (d-e): Ancestry was assigned based on proximity to 1KGP clusters. Participants received a primary ancestry label if their genetic profile matched a reference group with high confidence (Proportion ancestry >99% for EUR; >95% for AFR, CSA, and EAS). A more relaxed threshold was used for non-EUR ancestry groups due to smaller sample sizes, greater genetic diversity and to allow us to capture more participants within each group.

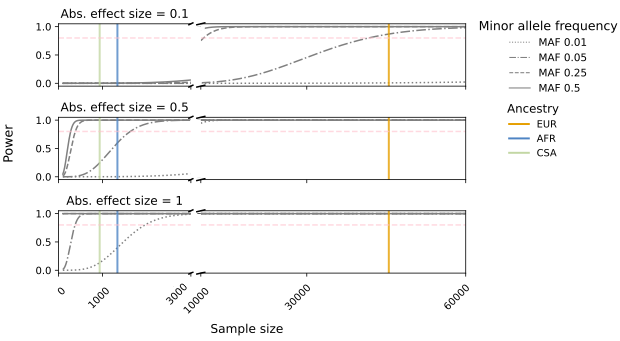

**Figure S2**: **Power calculations**. The figure displays the statistical power for detecting cis-vQTL associations at a significance threshold of p = 5x10^-8^ across a range of absolute effects sizes and minor allele frequencies, based on the framework of Sham and Purcell^1^. The dashed pink line indicates the 80% power threshold, with each ancestry group represented as a coloured vertical line: CSA (*N* = 934), AFR (*N* = 1,336) and EUR (*N* = 45,486).

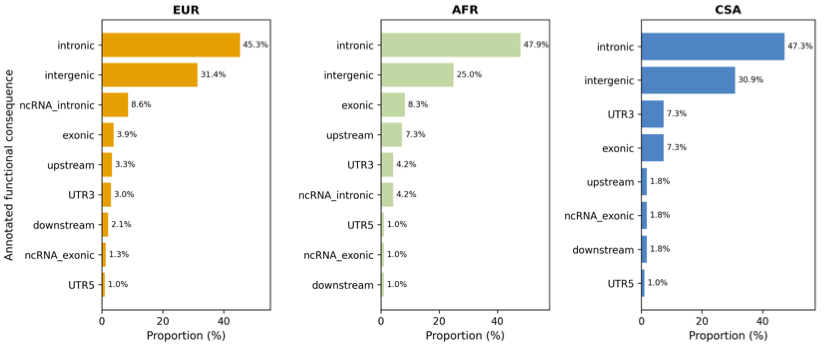

**d**

**c**

**b**

**a**

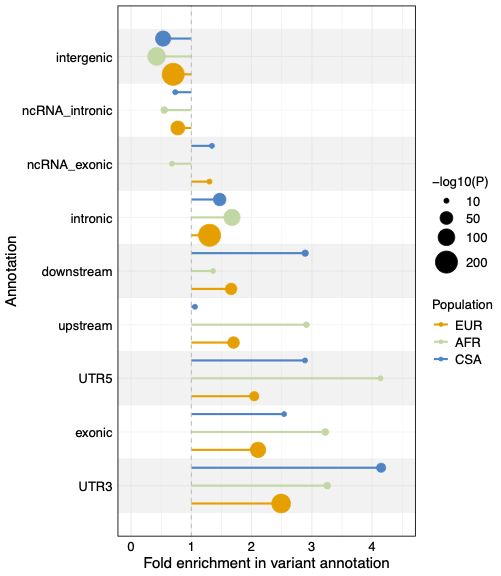

**Figure S3**: **Functional annotation of cis-vQTLs across ancestry groups**. Panels (**a-c**) Distributions of predicted functional annotation classes for all variance QTLs; bar height represents the mean proportion of variants within each class. (**d**) Fold enrichment of cis-vQTLs across different functional annotation classes, stratified by ancestry. For each annotation, lollipops represent the enrichment of cis-vQTLs relative to an ancestry-matched reference, with the size of the circle proportional to the significance. A fold enrichment greater than 1 indicates that the annotation is enriched among cis-vQTLs, values less than 1 indicate depletion. Values are capped (at -log10(P) = 200) for better visualization (see **Table S3**).

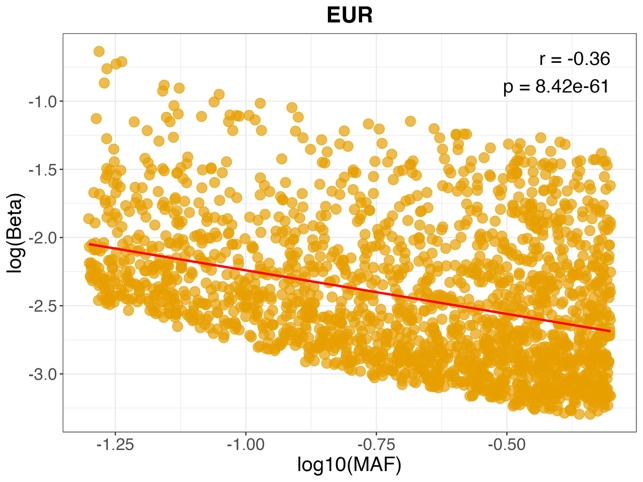

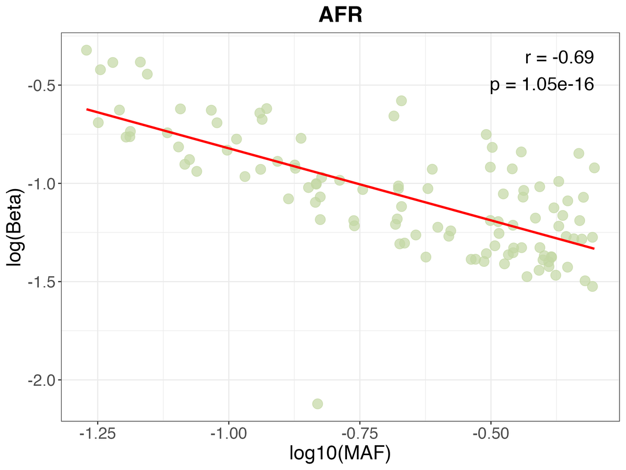

**c**

**b**

**a**

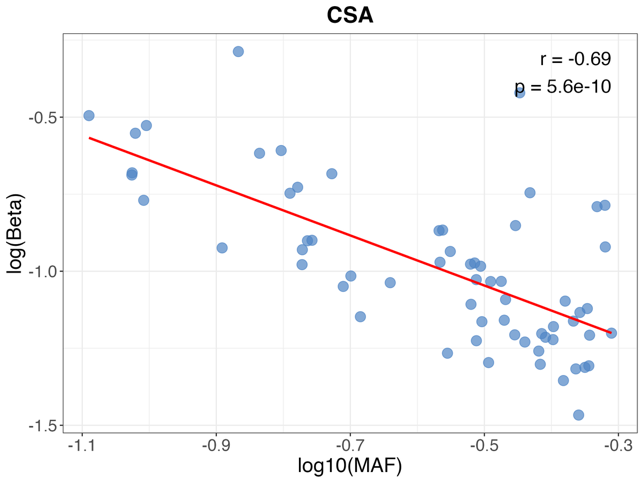

**Figure S4**: **Association between Effects sizes and MAF of vQTLs**. Panels (**a-c**): Relationship between the common logarithm of minor allele frequencies and the natural logarithm of absolute effect sizes for vQTLs across ancestries. The red lines represent the linear regression slopes for each population’s vQTL associations. P-values (unadjusted) were calculated using two-sided Pearson’s correlation tests on beta values for n = 104 (AFR), n = 61 (CSA) and n = 2,001 (EUR) cis-vQTL associations.

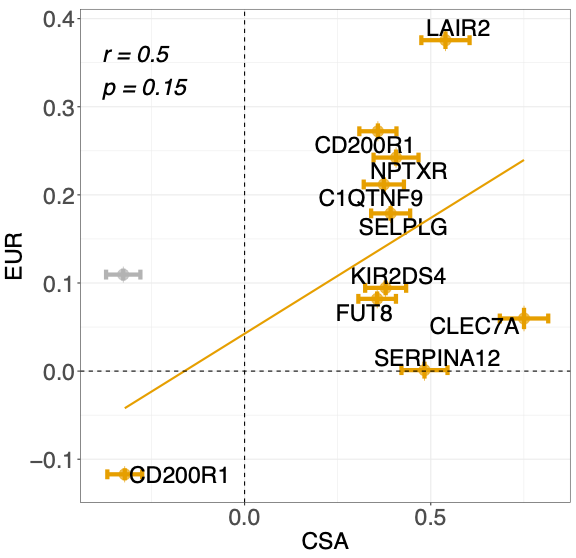

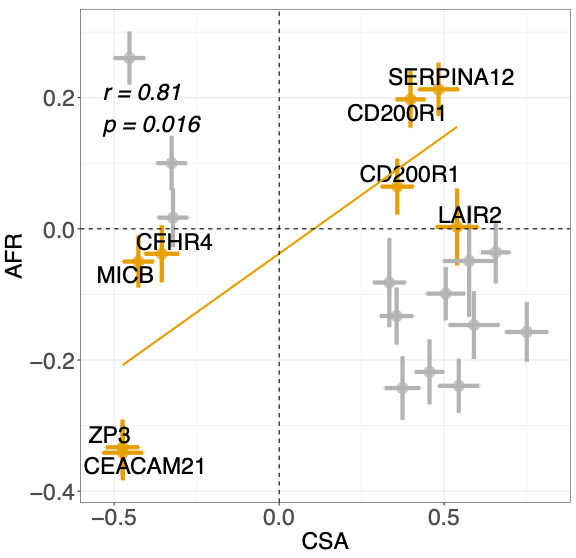

**b**

**a**

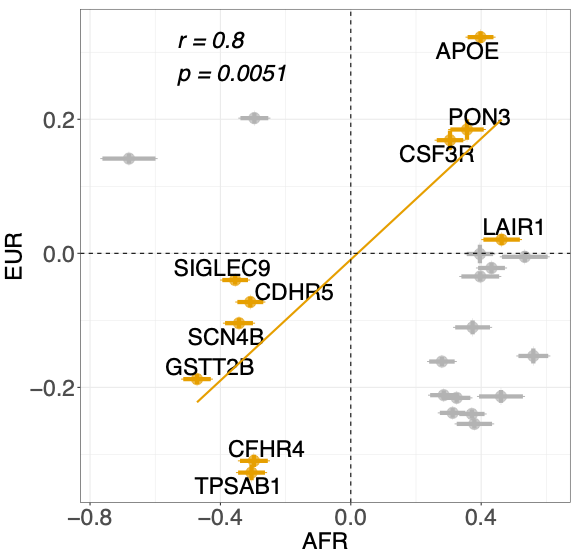

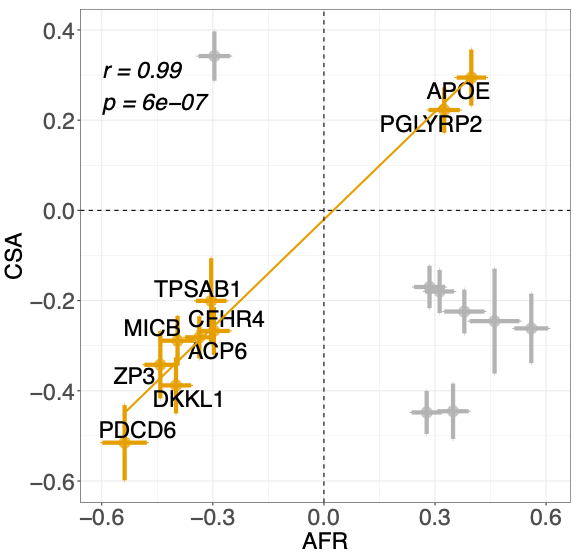

**d**

**c**

**Figure S5: Cross ancestry correlation analysis of cis-vQTL effect sizes**. We conducted pairwise comparisons of near-independent cis-vQTLs between (**a**) CSA vs EUR(**b**) CSA vs AFR; (c)AFR vs EUR; (d) AFR vs CSA ancestry pairs, to evaluate consistency in direction and magnitude of effect sizes. For each comparison, signals identified were protein matched and mapped to the second population (target, on the y-axis), excluding variants that failed QC, and restricted to those with MAF > 5% and P_VE_ < 0.05 in the target population. For each comparison, we present the correlation (R) in the top left corner.

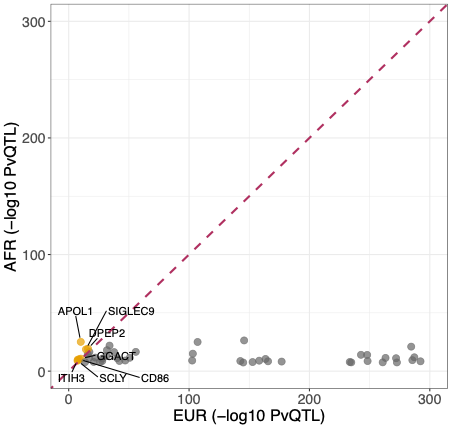

**Figure S6:** **Proteins with near independent-vQTLs in EUR and AFR only**. Proteins where the AFR -log10(P_VE_) is more significant than in EUR are annotated and highlighted in orange. P-values are capped (at -log10(P_VE_) = 300) for better visualization, excluding 27 proteins that were more significant in EUR. For proteins with multiple cis-vQTLs, only the most significant cis-vQTL is plotted. The dashed maroon line represents the equality line (x = y).

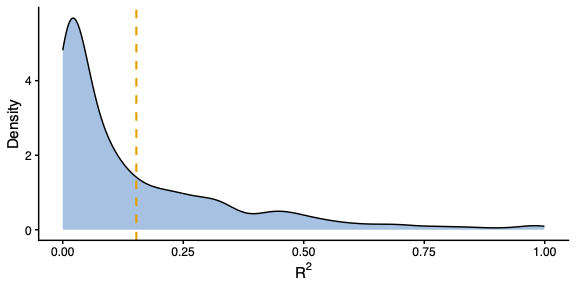

**Figure S7**. **Distribution of pairwise LD-r2 values between fine mapped cis-pQTLs from Sun et al 2023 and cis-vQTLs matched for the same protein**. For each protein, we plot the R² between each fine-mapped cis-pQTL and its highest pairwise LD vQTL proxy (excluding 45 exact matches). Orange dashed line indicates the mean R² = 0.18.

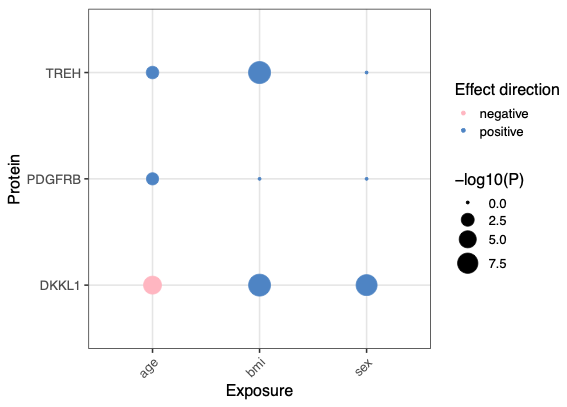

**Figure S8 Bubble plot displaying GxE interaction results for three exposures in EUR**. The plot highlights proteins with vQTLs significantly associated with age, sex, and BMI, with bubble colour representing the direction of effect and bubble size proportional to -log10 of FDR corrected p-values.

**a**

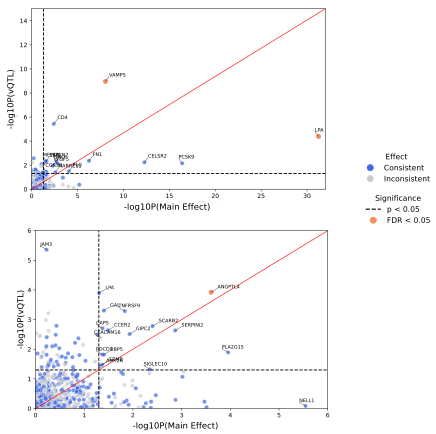

**b**

**Figure S9: Proteins with MVMR evidence for associations with CAD and T2D**. Panel (**a**) shows proteins with both main and variance (vQTL) effects associated with coronary artery disease (CAD), while panel (**b**) presents those associated with type 2 diabetes (T2D). Proteins showing consistent directions of effect for both main and variance effects are coloured blue, while those with inconsistent directions are shown in grey. Unadjusted p-values are shown in both panels. Black dashed lines indicate the nominal (p < 0.05) threshold and proteins are annotated when they pass threshold for both main and variance effects. Protein with significant p-values for both effects after FDR correction (FDR < 0.05) are highlighted in orange. In panel (b), two additional proteins with significant p-values for main effects (NELL1) and variance effects (JAM3) are also labelled despite not passing the p < 0.05 threshold for both components. The diagonal red line represents the equality line (x = y).

**a**

**b**

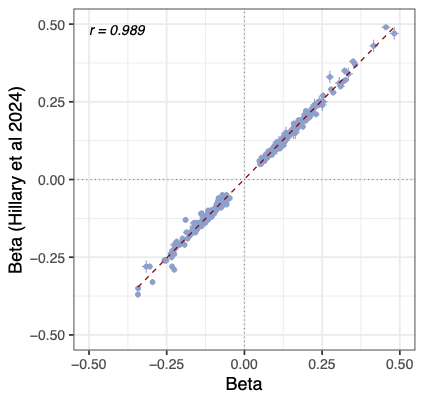

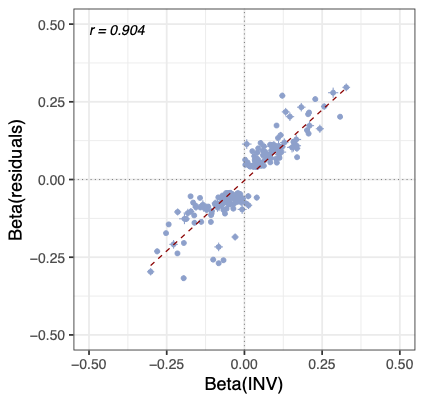

**Figure S10:** **Sensitivity analysis for vQTLs in EUR**. Panel (**a)** shows the correlation of effect sizes for 350 cis-vQTLs matched between Hillary et al, 2024. (shown on the y-axis) and this study. In both panels, the red dashed line represents the regression line, and the Pearson’s correlation coefficient (r) is indicated in the top-left corner. Panel (**b)** displays the correlation of effect sizes (Betas) for 100 proteins not assayed in the initial release of proteomic data for ~1500 proteins from the UKB. We compare the inverse normal transformed residuals (Beta (INV)) after covariate adjustment and untransformed residuals after covariate adjustment (Beta (residuals)).
